# Estimation of Total Immunity to SARS-CoV-2 in Texas

**DOI:** 10.1101/2021.08.05.21261610

**Authors:** Stacia M. Desantis, Luis G. León-Novelo, Michael D. Swartz, Ashraf S. Yaseen, Melissa A. Valerio, Frances A. Brito, Jessica A. Ross, Harold W. Kohl, Sarah E. Messiah, Steve H. Kelder, Leqing Wu, Shiming Zhang, Kimberly A. Aguillard, Michael O. Gonzalez, Onyinye S. Omega-Njemnob, Camille J. Breaux, David L Lakey, Jennifer A. Shuford, Stephen Pont, Eric D Boerwinkle

## Abstract

Given the underestimate of seroprevalence in the US due to insufficient testing, accurate estimates of population immunity to SARS-CoV-2 or vaccinations do not exist. Although model-based estimates have been proposed, they require inputting unknown parameters such as viral reproduction number, longevity of immune response, and other dynamic factors. In contrast to a model-based approach for estimating population immunity, or simplistic summing of natural- and vaccine-induced immunity, the current study presents a data-driven statistical procedure for estimating the total immunity rate in a region using prospectively collected serological data along with state-level vaccination data. We present a detailed procedure so that efforts can be replicated regionally to inform policy-making decisions relevant to SARS-CoV-2. Specifically, we conducted a prospective longitudinal statewide cohort serological survey with 10,482 participants and more than 14,000 blood samples beginning on September 30, 2020. Along with Department of State Health Services vaccination data, as of July 4, 2021, the estimated percentage of those with naturally occurring antibodies to SARS-CoV-2 in Texas is 35.3% (95% CI = (33.7%, 36.9%) and total estimated immunity is 69.1%. We conclude the seroprevalence of SARS-CoV-2 is 4 times higher than the state-confirmed COVID-19 cases (8.8%). This methodology is integral to pandemic preparedness.

## 1. Introduction

It is increasingly important to estimate the percentage of individuals in the US who may be protected from the novel SARS-CoV-2 virus as a result of having circulating anti-SARS-CoV-2 antibodies. People obtain immunity through either natural infection with SARS-CoV-2 or vaccination, and total immunity is the combination of these two avenues of immunity. Usually, an estimate of total immunity is obtained using mathematical modeling and simulation, which require inputs such as duration of immunity once infected, viral reproduction rate, population mixing, and additional factors [1, 2, 3, 4, 5]. However, the contributions of these inputs are still not fully known. For example, researchers are unsure of the duration of natural and vaccine-induced immunity, and possible T-cell cross-reactivity. Further, continual emergence of SARS-CoV-2 variants threaten progress toward immunity. [*e.g*., 6, 7, 8]

Recent research suggests neutralizing antibodies to SARS-CoV-2 persist for at least 5 months [9, 10] or possibly longer [11], and that re-infection risk is low in the several months after initial infection [12]. Additionally, at the time of this publication, there has been great success of mass vaccination’s in lowering viral transmission, *e.g*., [13] indicate that in Israel where approximately 61% of the population are vaccinated (>80% of adults). This resultant reduction in viral spread has inspired the idea of a path to normality [NYT, 14].

Given the above most current prevailing assumptions that: 1. Reinfection with COVID-19 within a few months is rare [15]; 2. Neutralizing antibodies from natural infections typically last at least 5 months and cross-reactivity of serological tests is rare [16, 17]; and, 3. Vaccination produces a robust and reasonably long-term antibody response, make it possible to estimate regional total immunity as a combination of natural and vaccine-induced immunity [18].

The goal of this report is to demonstrate this estimation process in Texas as of July 4, 2021 using a prospectively designed serological survey. To this end, we first estimate period seroprevalence over 1-week intervals from 14,899 blood specimens collected prospectively from participants throughout Texas. We then compute a census age-adjusted seroprevalence estimate of natural infection and combine it with the Texas Department of State Health Services (DSHS) de-identified population-level vaccination data to obtain an accurate state-level estimate of total immunity. Notably, the approach is not limited to the current pandemic; it is applicable to any infectious disease.

## 2. Methods

### 2.1. Participants and Study Design

The Texas Coronavirus Antibody REsponse Survey (Texas CARES) initiative has been previously described [19, 20]. Briefly, Texas CARES is a prospective convenience sample of adult retail/business employees, K-12 and university educators and university students, those attending Health Resources and Services Administration (HRSA)-designated federally qualified health centers (FQHCs), and children 5-17 years, all of whom agreed to longitudinal monitoring of SARS-CoV-2 antibody status every three months (three time points total) from 10/1/2020-9/30/2021. A consent form and survey questionnaire were administered online at each of the three time points. More details about the study are publicly available on the Texas CARES dashboard [21].

### 2.2. Serological Assay and Vaccination Records

Antibody status was determined using the Roche Elecsys^®^ Anti-SARS-CoV-2 (qualitative) assay detection of neutralizing antibodies against SARS-CoV-2 nucleocapsid (N) protein, hereafter referred to as “Roche N-test”. The test has a sensitivity (95% confidence interval, CI) of 99.5%(97.0,100.0) and specificity of 99.82%(99.69,99.91) >=14 days after infection. De-identified population level daily vaccination data (2 doses for mRNA vaccines or 1 dose for Johnson and Johnson vaccine) by age group were obtained from Texas Department of State Health Services (DSHS). All protocols were approved by the UTHealth Committee for the Protection of Human Subjects and were also deemed “public health practice” by the Texas DSHS IRB.

### 2.3. Statistical Methods

The following components are estimated from the data, (1) immunity from natural infection, (2) immunity from complete vaccination, and (3) total immunity defined as immunity from either natural infection or vaccination (*e.g*., “estimated total immunity”). Natural immunity period seroprevalence is calculated at a given time interval of Texas CARES, while vaccine-induced immunity is known (recorded by DSHS). For the current survey, a 1-week interval was deemed appropriate given the participant accrual rate into Texas CARES, and disease wave fluctuations. Within this interval, we assume serological status from prior infection and vaccine status are not independent events. This assumption is very well-supported by the data, which as of July 4th show 18.6% of those with reported prior documented COVID-19 disease are vaccinated, versus 30.7% without prior documented COVID-19. Our data and other research[22, 23] support that natural and vaccine-induced immunity are likely not independent events.

#### 2.3.1. Calculating Total Immunity to SARS-CoV-2 in Texas

We describe the methods used to compute the total immunity in Texas over time. Let *H* denote the total number of census age groups, *h* = 1, …, *H*. Assume we have serological and vaccination data for *T* weeks. Let *t* index the time window (week), where *t* = 1, …, *T*. Now define:

- *ν*_*ht*_: Vaccination proportion in age group *h* at week *t* (provided by state records). Since the vaccination status is cumulative, *ν*_*h*1_ *≤ν*_*h*2 …_ *≤ν*_*hT*_ for *h* = 1, …, *H*, and *ν*_*ht*_ is known with certainty.
- *η*_*ht*_: Natural immunity proportion in age group *h* at week *t*. This is unknown but is estimated cross-sectionally using the Roche N-test from Texas CARES.
- *w*_*h*_: Proportion of the Texas population in age group *h*. Thus, *w*_*h*_ is also known, and ∑_*h*_ *w*_*h*_ = 1.

The immunity rate in time window, *t*. (defined as having received the vaccine or testing positive for antibodies in the time window) in age group *h* is

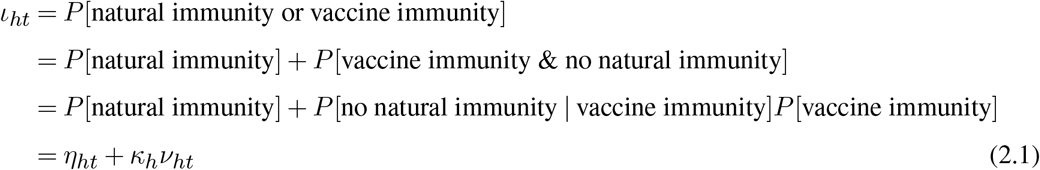

In (2.1) above, for brevity, we omit the text “in group age *h* at week *t*”. The definition of

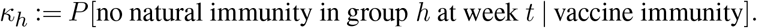

This implicitly assumes the probability is equal across all weeks, *t* = 1, …, *T*. Also notice that the proportion of the population with both natural and vaccine induced immunity is easily estimated as (1 *− κ*_*h*_)*ν*_*ht*_, so a mathematically equivalent expression to (2.1) is

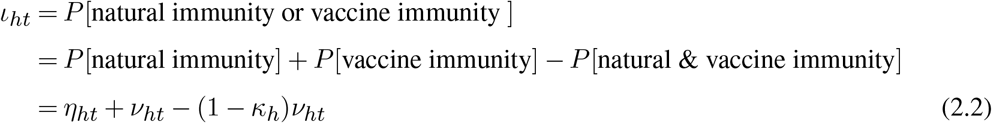

which may appear more intuitive: the total immunity rate is equal to the sum of the natural and vaccine induced immunity rates, minus their overlap. The population seroprevalence at week *t* is,

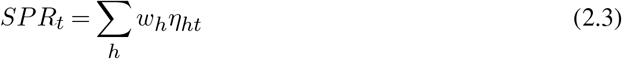

and the immunity rate (natural or vaccine induced) at week *t* in the population is,

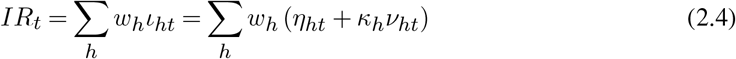

where *ι*_*ht*_ is given in (2.1). The population proportion with both natural and vaccine-induced immunity is,

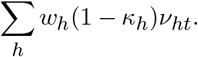

In order to estimate *SPR*_*t*_ and *IR*_*t*_ we must estimate *η*_*ht*_ and *κ*_*h*_. We show these steps in the following subsubsection. We also note that had we assumed independence between natural and vaccine induced immunity, *κ*_*h*_ = 1 *− η*_*ht*_, as expected.

#### 2.3.2. Estimation of Parameters for calculation of Total Immunity

We estimate *κ*_*h*_ and *η*_*ht*_ using the Roche N-test results. First, *κ*_*h*_ is estimated using the information from all *T* = 26 study weeks since January 1, 2021, as the following sample proportion,

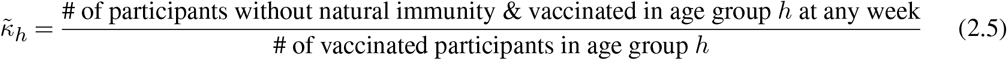

and *η*_*ht*_ is initially estimated as,

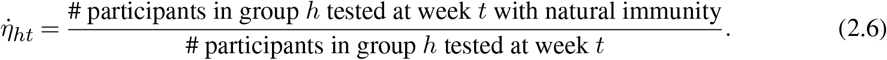

Once we have 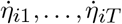 (and the denominators in the 2 equations above) we compute the isotonic version (across index *t*) of 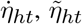 such that that 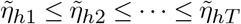 for *h* = 1, …, *H*. See Supplementary Materials Subsection S.1 for details of this calculation. The isotonic estimate of *η*_*ht*_ is appropriate here because it reflects the fact that seroprevalence should not decrease over a short time interval (even though its raw estimate 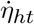 can decrease due to expected sampling error in a small window, *t*). Once these estimates are obtained, we compute the estimates of *SPR*_*t*_ and *IR*_*t*_, called 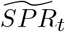 and 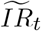, by substituting the values of 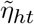 and 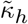 into equations (2.3) and (2.4). Construction of a 95% confidence interval for 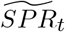 is based on that for a proportion from a stratified design in which the outcome variable is binary [*e.g*., 24, 25] (details provided in Supplementary Materials S.2).

#### 2.3.3. Algorithm to Estimate the Total Immunity Curve from Jan 1, 2021 to July 4, 2021

Recalling, *H* is the total number of age groups and *T* the total number of weeks. The algorithm is:

1. For, *h* = 1, …, *H*
  a. Using the Roche N-test, compute 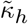 in (2.5).
  b. Obtain the (cumulative) state vaccination rate for week *t* by age group, denoted *ν*_*ht*_, from the Texas Dept of State Health Services database. Since they are cumulative, *ν*_*h*1_ *≤ ν*_*h*2_ *≤…≤ ν*_*hT*_ for *h* = 1, …, *H*.
  c. For *t* = 1, …, *T*, compute the preliminary estimated N-test positive rate in the study at week *t*, 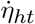 in (2.6). Next, compute the isotonic version of 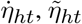, such that 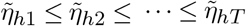
2. Estimate the age-adjusted seroprevalence rate at week *t*,

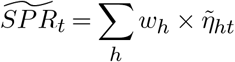

and then the total immunity rate at week *t*,

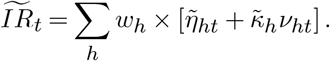
3. Plot *t v.s*. 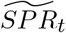 and *t v.s*. 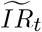

## 3. Results

Demographics of the full sample, and adults 18 years and over, respectively are shown in Tables 1 and 2. The mean (standard deviation) age of all participants was 45.9 years (16.1) and most participants were in the 50-54 year age group (30.9%). Most were female (69.4%), White (88.9%), and from urban locations (92.0%). Most adults reported having some college education or an advanced or professional degree, and were employed full time.

**Table 1:** Texas CARES participants’ demographics.

| N=10,482 |  |  |
| --- | --- | --- |
| **Age (years)** |  |  |
| Mean (SD) | 45.9 | (16.1) |
| Missing | 0 |  |
| **Age (categorical)** |  |  |
| 0-15 | 402 | (3.8%) |
| 16-17 | 124 | (1.2%) |
| 18-29 | 1257 | (12.0%) |
| 30-39 | 1802 | (17.2%) |
| 40-49 | 2331 | (22.2%) |
| 50-64 | 3244 | (30.9%) |
| 65-74 | 1100 | (10.5%) |
| 75-79 | 170 | (1.6%) |
| 80-84 | 39 | (0.4%) |
| 85+ | 13 | (0.1%) |
| Missing | 0 |  |
| **Gender** |  |  |
| Female | 7262 | (69.4%) |
| Male | 3208 | (30.6%) |
| None of these describe me | 1 | (0.0%) |
| Missing | 11 |  |
| **Race** |  |  |
| American Indian or Alaskan Native | 63 | (0.6%) |
| Asian | 470 | (4.7%) |
| Black | 382 | (3.8%) |
| Hawaiian or Other Pacific Islander | 18 | (0.2%) |
| Multi-racial | 192 | (1.9%) |
| White | 8974 | (88.9%) |
| Missing | 383 |  |
| **Ethnicity** |  |  |
| Hispanic | 2711 | (26.8%) |
| Non-Hispanic | 7409 | (73.2%) |
| Missing | 362 |  |
| **BMI (categorical)** |  |  |
| Underweight | 126 | (1.3%) |
| Normal | 3250 | (32.6%) |
| Overweight | 3131 | (31.4%) |
| Obese | 3458 | (34.7%) |
| Missing | 517 |  |
| **Geographic Location** |  |  |
| Rural | 824 | (8.0%) |
| Urban | 9488 | (92.0%) |
| Missing | 170 |  |

**Table 2:**
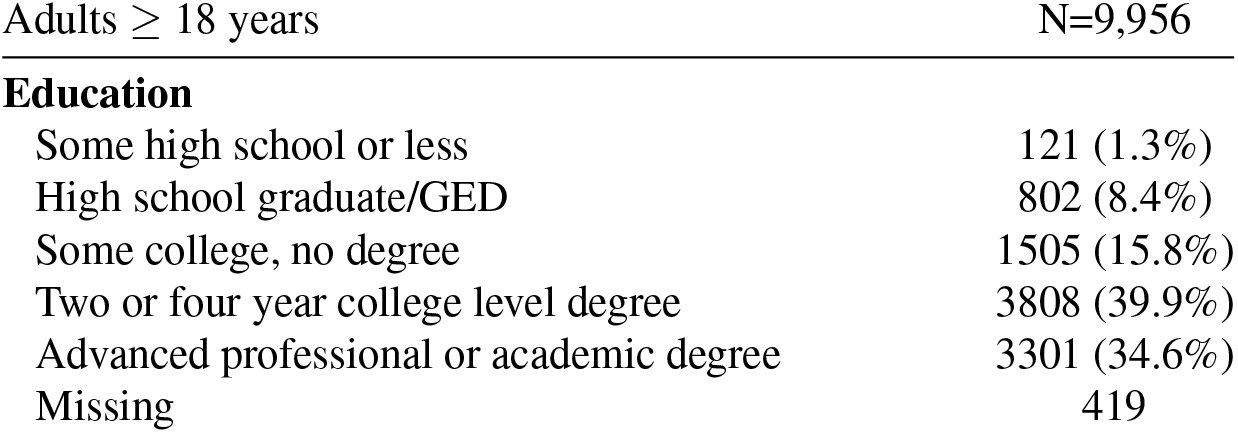
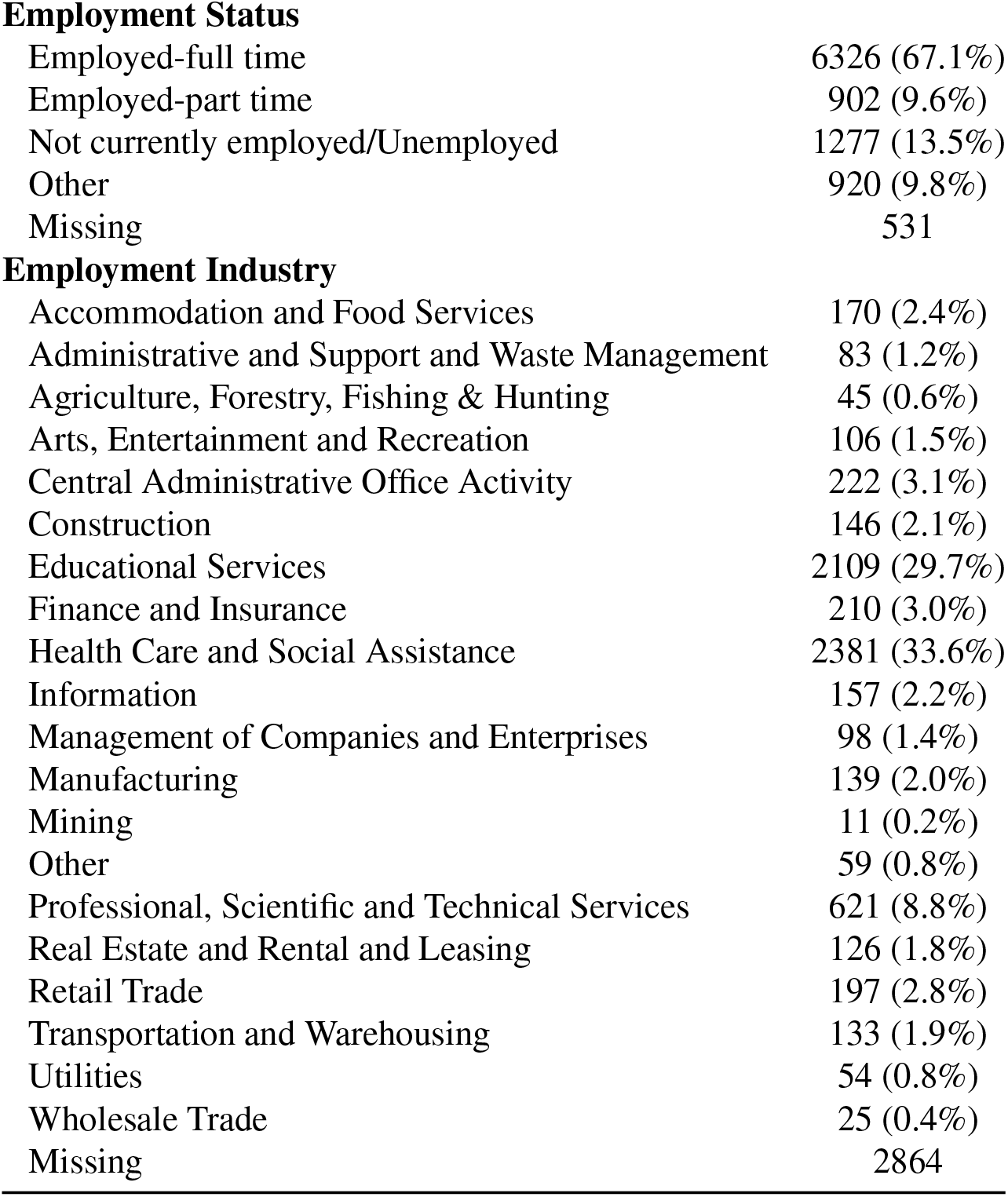
Texas CARES participants’ demographics for participants 18 and older.

|  |  |
| --- | --- |
| Adults ≥ 18 years | N=9,956 |
| **Education** |  |
| Some high school or less | 121 (1.3%) |
| High school graduate/GED | 802 (8.4%) |
| Some college, no degree | 1505 (15.8%) |
| Two or four year college level degree | 3808 (39.9%) |
| Advanced professional or academic degree | 3301 (34.6%) |
| Missing | 419 |
| **Employment Status** |  |
| Employed-full time | 6326 (67.1%) |
| Employed-part time | 902 (9.6%) |
| Not currently employed/Unemployed | 1277 (13.5%) |
| Other | 920 (9.8%) |
| Missing | 531 |
| **Employment Industry** |  |
| Accommodation and Food Services | 170 (2.4%) |
| Administrative and Support and Waste Management | 83 (1.2%) |
| Agriculture, Forestry, Fishing & Hunting | 45 (0.6%) |
| Arts, Entertainment and Recreation | 106 (1.5%) |
| Central Administrative Office Activity | 222 (3.1%) |
| Construction | 146 (2.1%) |
| Educational Services | 2109 (29.7%) |
| Finance and Insurance | 210 (3.0%) |
| Health Care and Social Assistance | 2381 (33.6%) |
| Information | 157 (2.2%) |
| Management of Companies and Enterprises | 98 (1.4%) |
| Manufacturing | 139 (2.0%) |
| Mining | 11 (0.2%) |
| Other | 59 (0.8%) |
| Professional, Scientific and Technical Services | 621 (8.8%) |
| Real Estate and Rental and Leasing | 126 (1.8%) |
| Retail Trade | 197 (2.8%) |
| Transportation and Warehousing | 133 (1.9%) |
| Utilities | 54 (0.8%) |
| Wholesale Trade | 25 (0.4%) |
| Missing | 2864 |

We applied the method with *H* = 10 age groups, 0-15, 16-17, 18-29, 30-39, 40-49, 50-64, 65-74, 75-79, 80-84 and 85+ years. The census age-adjusted Texas COVID-19 seroprevalence using the Roche N-protein test over time (*i.e*., *t v.s* 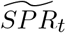) along with the 95% confidence band is shown in Figure 1. The vertical line on the graph delineates the time of first vaccine availability. The surges in seroprevalence correspond well to the known waves of SARS-CoV-2 in Texas [26].

**Figure 1.**
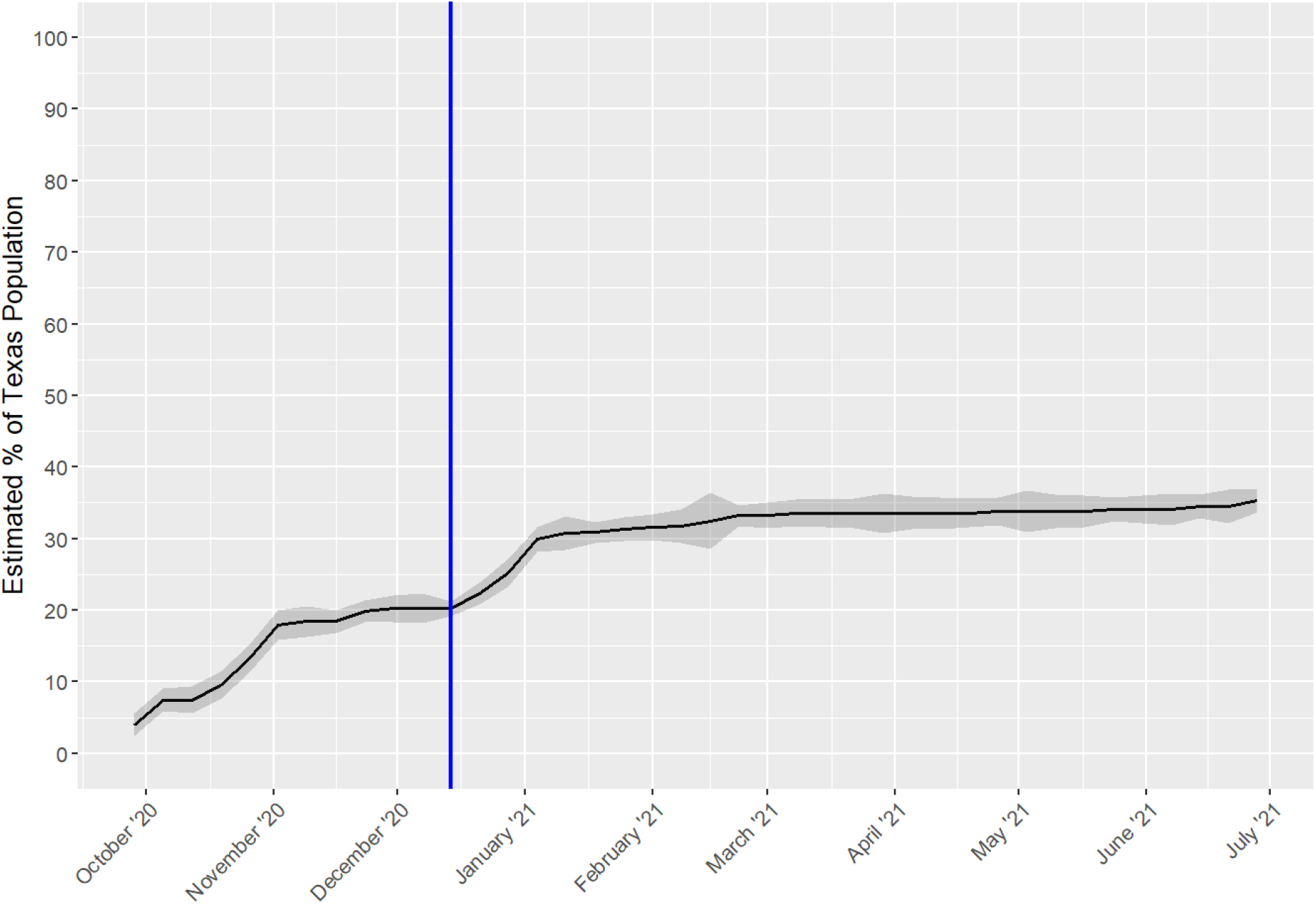
Weekly Natural Immunity Texas Cares Roche N-Test. Horizontal axis labels denote the first day of the month.

We note that 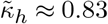 in all age groups; thus, the proportion of study participants who reported having had both COVID-19, and being fully vaccinated were roughly 1 −*κ*_*h*_ *≈* 17%. This indicates a violation of independence of natural infection and vaccination, which was expected. As these people must not be counted twice in the total immunity estimate, they are subtracted appropriately in each time period (week) per Equation (2.2).

The estimated age-adjusted total period immunity in Texas, defined as immunity from either natural infection or full vaccination (solid line) over time (*i.e*., *t v.s* 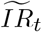) is shown in Figure 2. As of July 4, 2021, total immunity is estimated at 69% of the Texas population, with approximately 35.3% (95% CI = (33.7, 36.9) resulting from natural infection. To our knowledge, this is the most robust and accurate non-model based estimate of total immunity to date in the state of Texas.

**Figure 2.**
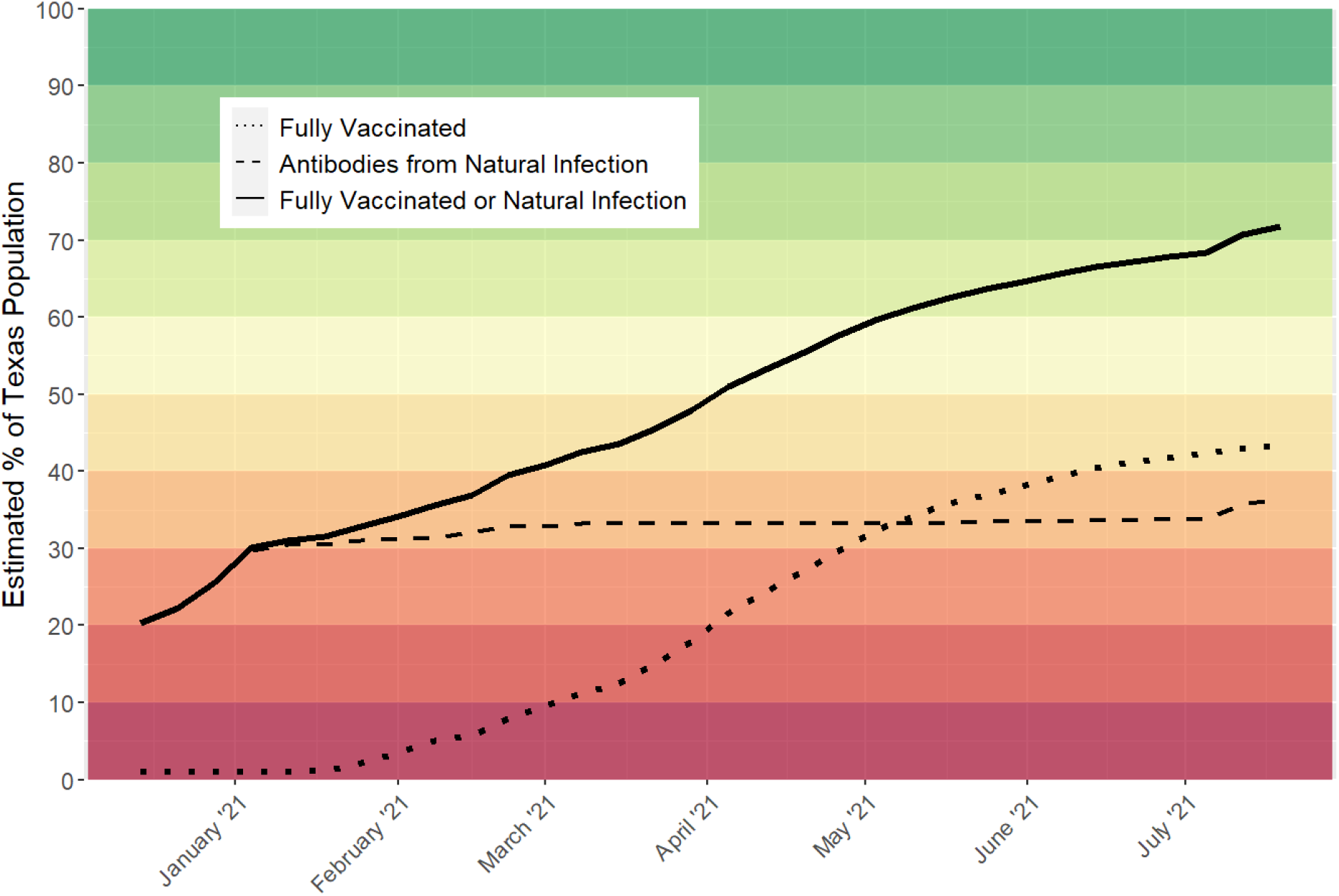
Estimated total immunity in Texas (i.e., weekly percentage of fully vaccinated or naturally occurring antibodies). Horizontal axis labels denote the first day of the month. The estimate as of July 4, 2021 is 69.08%.

We do not include a confidence interval for total immunity since the proportion vaccinated is a known (fixed) population quantity rather than an estimate, and thus does not lend itself to an estimate of variability. While the seroprevalence is not known or fixed, the large sample of 14,899 blood specimens would result in a very small range for the 95% confidence interval, if one were to be produced for the total immunity line.

## 4. Discussion

Using the methods proposed, the estimated proportion of the Texas population with antibodies against the SARS-CoV-2 virus, either from natural infection or induced by the vaccine, is nearly 70% as of July 4, 2021. This means 70% of the population benefit from some degree of protection from reinfection from SARS-CoV-2 and acquiring COVID-19. There are several challenges to further practical or applied interpretation of these data. First, we do not know the relative degree of protection from antibodies from a natural infection compared to antibodies from the vaccine. The titer of antibodies from a full vaccine regimen is higher than a typical natural infection [27], but the diverse epitopes of a natural infection may offer advantages over antibodies targeting only spike protein. In addition, the SARS-CoV-2 mutates producing new strains that will likely influence the degree of protection of circulating antibodies [28]. Second, there is limited data on how long antibodies to the vaccine and to natural infection last [*e.g*., 29, 30, 31, 32, 33] Though early findings about the duration of natural and vaccine-generated immunity are promising, it is reasonable to expect the proportion of people with detectable antibodies will decline over time. And finally, protection from an infectious agent is complex, and the concept of seroprevalence and protection does not take into account cell-mediated immunity and physical barriers, such as masks.

In contrast to model-based approaches, the current research will allow researchers and health departments to calculate regional estimates of total immunity in the least biased manner.

Limitations may occur in observational serological surveys; e.g., sample demographics may not be fully representative of the state, which is true for some variables in the current survey. Sampling variability or selection biases may operate within small time windows of a serological survey, and can result in inaccuracies in seroprevalence estimates. It will therefore generally be necessary to smooth estimates using a chosen time window dependent on factors such as the magnitude of the wave of infection and participant accrual rate. Fortunately, we observe that the application of an isotonic restriction to reflect the assumption that seroprevalence should not decrease in a reasonably small time window mostly overcomes the issue of daily or weekly sampling variability. Further, it is necessary to estimate the percentage of people who have both had natural COVID-19 infection and are fully vaccinated in a given time window in order to subtract that proportion from the overall sum. Finally, it is important to age-adjust estimated serological and vaccination rates to the state census so they are commensurate with population demographics. This is especially important since vaccination was rolled out by age group, with older adults first priority in January-March 2021.

To our knowledge, this is the first fully data-driven estimation of total immunity to SARS-CoV-2 in the state of Texas, which is the second largest state in the US with a population of 29.2 million. The method proposed and applied here can be applied to any state or geographic area using vaccine counts, and an estimate of seroprevalence. As the pandemic unfolds and new variants are introduced, immunity will require further investigation and re-estimation.

## Data Availability

The data are in process of being collected over a period of 2 years. However summaries of the data are publicly available within the referenced URLs in the paper (the Texas CARES public dashboard: https://sph.uth.edu/projects/texascares/dashboard )

## Supplementary Material

### S.1. Order Restricted (Isotonic) Estimation of Probabilities

The algorithm below is retrieved from Algorithm 3 in https://core.ac.uk/download/pdf/33107977.pdf. The maximum likelihood estimate of *π* = (*π*_1_, *π*_2_, *…, π*_*H*_) under the restriction of *π*_1_ *≤ π*_2_ *≤ …≤ π*_*H*_, is calculated in the following way: Let *n*_*h*_ the number of observations in group *h*,

Step 1: Do 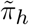 equal to the sample proportion in group *h*.

Step 2: While not *π*_*h*_ *≤ π*_*h*+1_, for *h* = 1,, *H −* 1, do

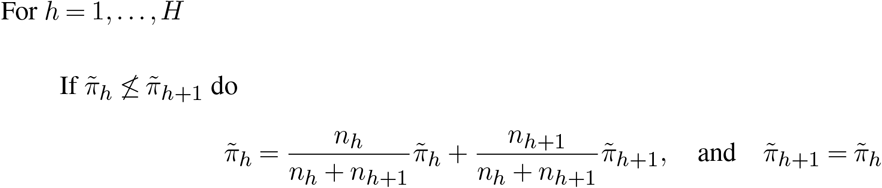

### S.2. Confidence Interval for the Seroprevalence

The construction of the confidence interval for 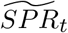 is based on the confidence interval for a proportion under a stratified sampling design [*e.g*., 24, 25]. Recall, the weight *w*_*h*_ = *N*_*h*_*/N* and *N*_*h*_ denote the proportion of and the number of individuals in the population in the age group *h*, respectively, and *N* the population total. An estimate of the sampling variance of the sample proportion 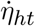 is

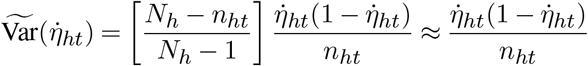

where *n*_*ht*_ is the number of participants in age group *h* (at week *t*). The approximation in the above equation is valid since *N*_*h*_ *>> n*_*ht*_. We base the confidence interval on this equation plugging in 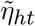 instead of 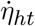

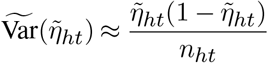

Then the sampling variance of 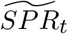 is estimated with

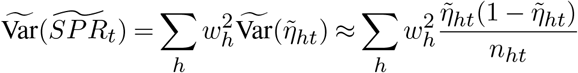

The asymptotic 1 *− α* confidence interval for *SPR*_*t*_ is then

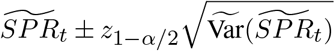

where *z*_*α*_ is the percentile *α* of the standard normal distribution, so when 1 *− α* = 0.95, *z*_1*−α/*2_ = 1.96.

